# Stratification of Systemic Lupus Erythematosus Patients Using Gene Expression Data to Reveal Expression of Distinct Immune Pathways

**DOI:** 10.1101/2020.08.25.20181578

**Authors:** Aditi Deokar

## Abstract

Systemic lupus erythematosus (SLE) is the tenth leading cause of death in females 15-24 years old in the US. The diversity of symptoms and immune pathways expressed in SLE patients causes difficulties in treating SLE as well as in new clinical trials. This study used unsupervised learning on gene expression data from adult SLE patients to separate patients into clusters. The dimensionality of the gene expression data was reduced by three separate methods (PCA, UMAP, and a simple linear autoencoder) and the results from each of these methods were used to separate patients into six clusters with k-means clustering.

The clusters revealed three separate immune pathways in the SLE patients that caused SLE. These pathways were: (1) high interferon levels, (2) high autoantibody levels, and (3) dysregulation of the mitochondrial apoptosis pathway. The first two pathways have been extensively studied in SLE. However, mitochondrial apoptosis has not been investigated before to the best of our knowledge as a standalone cause of SLE, independent of autoantibody production, indicating that mitochondrial proteins could lead to a new set of therapeutic targets for SLE in future research.

## 1. Introduction

Systemic lupus erythematosus (SLE) is the tenth most common cause of death among females 15-24 years old in the US (Yen and Singh, 2018). SLE is one of many autoimmune diseases, which are diseases in which a patient’s immune system mistakes parts of their own body as foreign, attacking their healthy organs and tissue (Lupus Foundation of America, 2020).

SLE can be driven by defects in the innate immune system and/or the adaptive immune system. SLE patients are often characterized by high levels of interferon-1, which causes inflammation in the innate immune system in response to viruses. In SLE, high interferon levels can be caused by a variety of factors, such as neutrophil extracellular traps (Bengtsson and Rönnblom, 2017). Most SLE patients also have high levels of autoantibodies, which are antibodies directed against self cells and are created by mature B cells (plasma cells) (Dema and Charles, 2016). Autoantibodies cause a much more targeted response than the innate immune system, but SLE patients can have a wide range of autoantibodies - one study found over 180 autoantibodies expressed in SLE patients (Yaniv et al., 2015). Some patients with lupus do not even have autoantibodies, and many of the autoantibodies in SLE are also found in other rheumatic diseases (Egner, 2000).

The heterogeneity of lupus symptoms and immune pathways affected makes it difficult to treat, because different drugs work well on different patients. Merrill et al. (2017) found that certain standard drugs (anti-rheumatic drugs and immunosuppressants) affect immune pathways differently in interferon-low and interferon-high patients. While there is still debate on whether SLE is one disease or many (Agmon-Levin et al., 2012), it is clear that subdividing SLE patients into categories will help treat patients.

Previous studies have tackled this problem by dividing patients based on antibody levels (Artim-Esen et al., 2014), gene expression (Toro-Domínguez et al., 2018), and immune molecule levels (Hamilton et al., 2018). However, none of these studies have reached a consensus on the best subdivision of SLE. Guthridge et al. (2020) used all three of these factors to divide SLE patients into seven clusters with unsupervised machine learning. But, in practice, gathering these different types of patient data to categorize a patient into an SLE subdivision is infeasible.

In this study, we use unsupervised machine learning to categorize SLE patients using only gene expression data, which is more accessible than all three types of data combined. This would help determine if gene expression data alone reveals similar patterns in immune pathway expression as does its combination with antibody levels and immune molecule levels.

## 2. Data and Methods

We use a gene expression dataset available on GEO (accession number GSE138458) containing data collected by Guthridge et al. (2020). The data includes 336 samples in total, with 24 control patients and 198 SLE patients. 108 of the SLE patients have two or more samples taken. Data pre-normalized by Guthridge et al. (2020) is used, employing bgAdjust background correction, vst variance stabilizing transformation, and rank invariant normalization, and outlier removal (1 control and 5 SLE).

Given the high dimensionality nature of gene expression data, dimension reduction techniques are required. Prior work has used unsupervised learning following dimension reduction of gene expression data for other diseases such as cancer (Shi and Luo, 2010). The dimensionality of our 47,323 gene data is reduced using three separate methods: *Principal Component Analysis* (PCA), *Uniform Manifold Approximation and Projection* (UMAP), and a simple *autoencoder* (AE), which are intended to minimize the effects of random variation on the unsupervised clustering model in different ways.

With both PCA and UMAP, 200 reduced features are selected. In PCA, these explain 96.29% of the variance in the original 47,323 genes. UMAP is a nonlinear model (unlike PCA) similar to t-SNE, used for visualization as well as nonlinear dimension reduction (McInnes et al., 2020). The autoencoder aims to reduce the loss of information between the original inputs (genes) and the decoded output of the same dimension. AEs with both linear and sigmoid activation functions are validated and the linear AE is found to perform much better after 100 epochs (validation loss 0.068) than the sigmoid AE (validation loss 48.28). So, we select the 1000 encoded components from the linear AE for subsequent clustering.

The three datasets with reduced features are then used for k-means clustering. To determine the best number of clusters, we use distortion score, silhouette score, and Calinski-Harabasz score. All these metrics are found to converge on 6 clusters for each dataset. k-means clustering is then used to derive 6 clusters from each dataset.

For visualization and interpretation of the clusters, we use 27 pre-existing modules created by Chaussabel et al. (2008). Each module represents a group of genes with a common function. These are used to calculate module scores for the three datasets. Module scores for each cluster represent the percentage of genes in each module that were significantly upregulated (i.e., overexpressed) or downregulated (i.e., underexpressed) in that cluster as compared to the controls, based on a two-tailed t-test (*p <* 0.05).

## 3. Results and Discussion

Figure 1 shows heatmaps generated from the module scores for the 3 feature-reduced datasets. These heatmaps show the percentage of underexpressed (brown) or overexpressed (purple) genes for SLE patients as compared to the controls.

**Figure 1:**
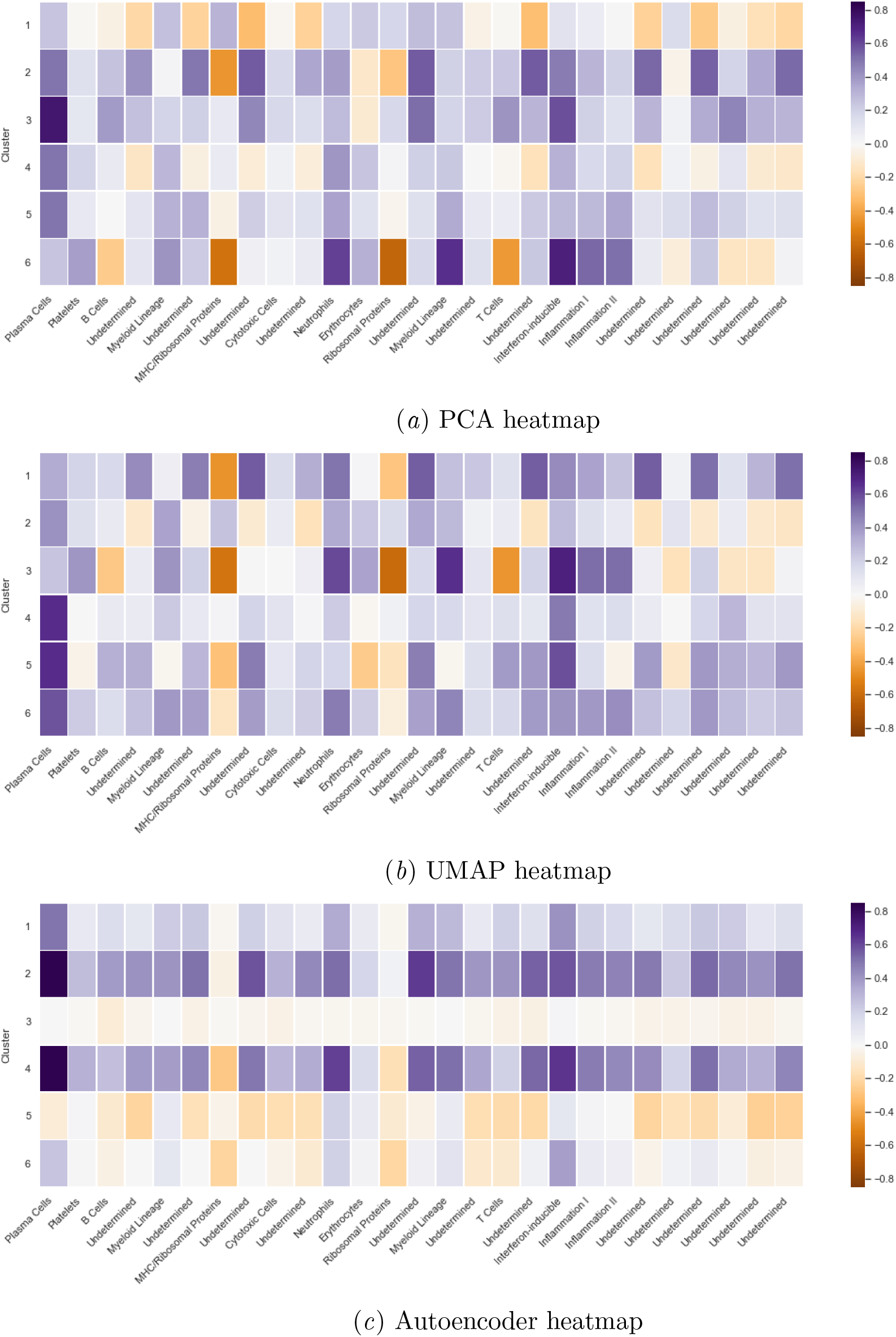
Comparative gene underexpression or overexpression for gene expression modules across clusters.

The clusters originating from the PCA and UMAP dimensionality reductions show very similar patterns in the upregulated and downregulated modules, while the clusters originating from the AE mostly show a consistent level of increased or decreased gene expression across all modules, excepting cluster 5.

The patients in the clusters created from the PCA and UMAP dimensionality reduction techniques and AE cluster 5 can be designated as belonging to one of three groups: (a) interferon-driven SLE, (b) autoantibody-driven SLE, and (c) SLE caused by mitochondrial apoptosis. The first two groups substantiate results from prior literature, while the third group presents a pathway that suggests a novel cause of SLE.

### 3.1 Interferon-driven SLE

In lupus, type 1 interferon levels are often elevated, which can lead to inflammation and tissue damage caused by the innate immune system (Crow, 2014). PCA cluster 6 and UMAP cluster 3 in Figure 1 both display substantial upregulation of genes related to interferons and inflammation. These two clusters validate the patterns also observed in Guthridge et al. (2020)’s clusters 1, 4 and 6. All of these upregulated genes are related to the innate immune response. Those patients also have underexpressed B and T cells and normal expression of plasma cells, which would all be overexpressed if production of autoantibodies by plasma cells was the main reason for autoimmunity, rather than interferon levels.

### 3.2 Autoantibody-driven SLE

Many of the other PCA and UMAP clusters displayed upregulation of antibody-producing plasma cells; particularly PCA clusters 2, 3, 4 and 5 and UMAP clusters 4, 5, and 6. Guthridge et al. (2020) observed a similar trend, where their clusters 2, 3, and 5 had higher T cell, B cell, and plasma cell related expression. While autoantibodies are known to be common in SLE, the diversity of autoantibodies (as discussed in Yaniv et al. (2015)) means that there is still work to be done understanding what is different among the antibodies produced in these four PCA clusters and three UMAP clusters. Some of these differences might come from genes used to create the clusters that were not included in the modules used for the heatmap visualization.

Brant et al. (2020), who grouped lupus patients based on their correlation between gene expression and disease activity, found one cluster where neutrophil levels correlated to disease activity and one where lymphocyte levels correlated to disease activity. Since neutrophil extracellular traps are one way that interferon levels become elevated, their neutrophil-correlated group might correspond to our high-interferon group, and their lymphocyte-correlated group might correspond to our antibody-driven group. More analysis should be done on disease activity correlation in our data to confirm this.

### 3.3 SLE caused by mitochondrial apoptosis

PCA clusters 1 and 4, UMAP cluster 2, and Autoencoder cluster 5 display a different pattern from many of the other clusters. In these clusters, many of the modules labeled as *Undetermined* by Chaussabel et al. (2008) were underexpressed. A closer look at the genes in these *Undetermined* modules reveals that they include mitochondrial ribosomal proteins, mitochondrial elongation factors, and proteins in the cAMP-signaling pathway. Mitochondrial ribosomal proteins, in addition to their ribosomal functions, are involved in apoptotic (programmed cell death) pathways (Kim et al., 2017), and cAMP signaling regulates mitochondrial apoptosis (Valsecchi et al., 2013). Apoptosis is known to be a factor in SLE, but mainly because ineffective clearance of apoptotic cells can expose B and T cells to intracellular material, leading to the creation of autoantibodies against this intracellular material (Mevorach, 2003).

We suggest that for the patients in these clusters, dysregulation of mitochondrial pathways or signaling from outside molecules, possibly lymphocytes, could cause mitochondrial apoptotic pathways to become activated in healthy cells, destroying healthy cells as is characteristic of SLE. These healthy cells would have a range of gene expression of mitochondrial proteins, but the cells with higher expression of the proteins would activate the apoptotic pathway and die. Only cells with lower expression levels would survive, so lower expression levels were found in our study. These lower expression levels would also impair mitochondrial functions, which has been observed to be true in SLE patients (Leishangthem et al., 2016).

Cluster 7 from the Guthridge et al. (2020) study also had low expression of mitochondrial respiration and mitochondrial stress genes (not discussed in their study). The discovery of this cluster of patients using two completely different machine learning approaches corroborates the idea that the mitochondrial apoptotic pathway is a novel cause for SLE. Future studies should investigate to a further extent the mitochondrial apoptotic pathway in SLE patients as a reason for destruction of self cells in addition to a way that autoantibodies are produced.

## 4. Conclusion and Future Work

In this study, we separated SLE patients into clusters based on their gene expression data using unsupervised learning. The data was collected by Guthridge et al. (2020), who clustered patients using antibody levels and immune phenotyping in addition to gene expression levels. We used only gene expression data and used entirely different methods from their study, in order to determine whether we would find similar clusters of patients. The dimensionality of the gene expression data was first reduced by three separate methods (PCA, UMAP, and a simple linear autoencoder) and the results from each of these methods were used to separate patients into six clusters with k-means clustering. These clusters revealed there were three separate immune pathways in the SLE patients causing SLE. These pathways were 1) high interferon levels, 2) high autoantibody levels, and 3) dysregulation of the mitochondrial apoptosis pathway. All three of these pathways were present in Guthridge et al. (2020)’s clusters, but to our knowledge this study is the first to propose mitochondrial apoptosis as a standalone cause of SLE, independent of autoantibody production. Future studies should investigate to a further extent the mitochondrial apoptotic pathway in SLE patients as a reason for destruction of self cells in addition to a way that autoantibodies are produced and investigate mitochondrial proteins as possible therapeutic targets for SLE.

## Data Availability

All data used in this study are available on GEO (accession number GSE138458) and were originally collected and referenced in the following study.
Guthridge, J. M., Lu, R., Tran, L. T. H., Arriens, C., Aberle, T., Kamp, S., Munroe, M. E., Dominguez, N., Gross, T., DeJager, W., Macwana, S. R., Bourn, R. L., Apel, S., Thanou, A., Chen, H., Chakravarty, E. F., Merrill, J. T., & James, J. A. (2020). Adults with systemic lupus exhibit distinct molecular phenotypes in a cross-sectional study. EClinicalMedicine, 20, 100291. https://doi.org/10.1016/j.eclinm.2020.100291

https://www.ncbi.nlm.nih.gov/geo/query/acc.cgi?acc=GSE138458

## Appendix A. Analyzing Patients with Multiple Samples

For patients who had multiple samples taken, k-means following the autoencoder classified them into the same cluster 97.3% of the time, while k-means following PCA and UMAP classified them into the same cluster 32.9% and 45.7% of the time respectively (Figure 2). While gene expression data is correlated with SLE disease activity (Kegerreis et al., 2019; Toro-Domínguez et al., 2018), Petri et al. (2019) found that the majority of gene expression signatures were stable in patients over time. This suggests that the autoencoder’s dimensionality reduction may have emphasized the stable gene expression signatures, causing them to be a major factor in the clustering, but that PCA and UMAP, which aimed to preserve more of the variance in the data, did not maintain the data from genes whose expression was stable over time. Many of these more stable genes might not have been related to the immune system, so they were not included in Chaussabel et al. (2008)’s coexpression modules. Thus, the more variable modules that were in the heatmap would have shown a lot of variation between the patients in each cluster, causing the clusters in the autoencoder heatmap to show a more consistent level of expression across all genes in the modules. Further analysis should be done to determine the level of variation in gene expression in the modules for the autoencoder clusters in comparison to the PCA and UMAP clusters, and to determine whether the PCA and UMAP clusters correlated to disease activity more than the autoencoder clusters, which this idea would imply.

**Figure 2:**
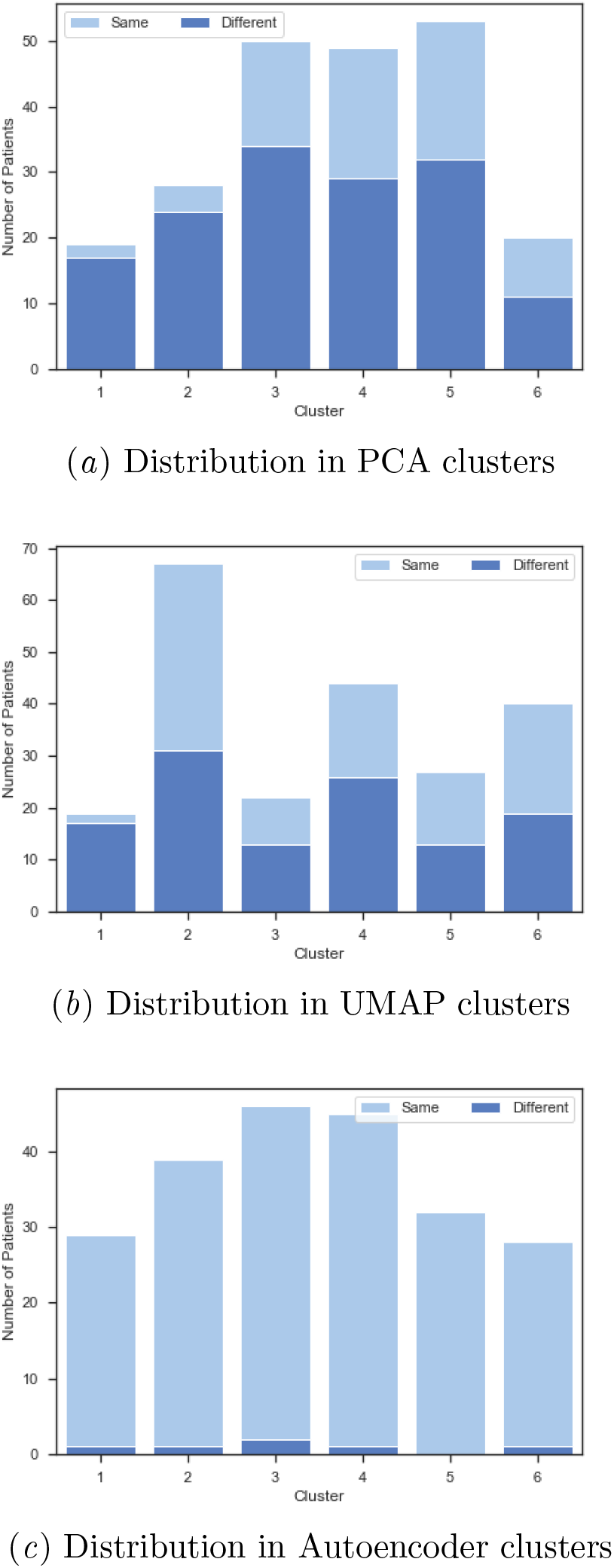
Patients in each cluster, of those who had multiple samples taken, who were put in the same cluster or different cluster for (a) PCA clusters, (b) UMAP clusters, (c) Autoencoder clusters.

## Appendix B. Dimensionality Reduction Models’ Specifications

The following specifications were used in the three dimensionality reduction techniques.

- *Autoencoder* : `keras` API was used for the simple autoencoder with: Input dimension: `(47323,)`, Output dimension: `(1000,)`, `activation = ‘linear’` in encoded and decoded layers, `epochs = 50`, `optimizer = ‘adam’`, `loss = ‘mse’`, `batch_size = 64`, `shuffle = True`, `validation_split = 0.2`.
- *UMAP* : `UMAP` parameters used: `n_components = 200`, `n_neighbors = 15`, `min_dist = 0.1`, `metric = ‘euclidean’`.
- *PCA*: Linear dimensionality reduction using Singular Value Decomposition (SVD) was used with the PCA class in `sklearn` API with the following parameters: `n_components = 200`, `svd_solver = ‘randomized’`.

